# Changes in Apolipoprotein A1 and B, Glucose Metabolism, and Skeletal Muscle Mass in Peripheral Artery Disease after Endovascular Treatment: A Pilot Study

**DOI:** 10.1101/2022.04.01.22273293

**Authors:** Takeshi Ikeda, Hidenori Komiyama, Tomoyo Miyakuni, Masamichi Takano, Masato Matsushita, Nobuaki Kobayashi, Kuniya Asai, Wataru Shimizu, Yasushi Miyauchi

## Abstract

Peripheral artery disease impairs walking and physical activity, resulting in further loss of skeletal muscle. However, peripheral artery disease can be treated with endovascular treatment. The thigh muscle has been shown to correlate with systemic skeletal muscle volume. We assessed the changes in the metabolism of apolipoproteins A1 and B, blood glucose, and thigh muscle mass after endovascular treatment in above-the-knee peripheral artery disease where vessels supply the thigh muscle. Mid-thigh muscle volume was measured with computed tomography before and at 6 months after endovascular treatment. Apolipoproteins A1 and B, fasting blood glucose, post-load (75 g oral glucose tolerance test) 2 h-blood glucose, and glycated hemoglobin A1c (HbA1c) levels were measured concomitantly. The relationships between changes in apolipoproteins A1, apolipoproteins B, blood glucose, post-oral glucose tolerance test 2 h-blood glucose, Rutherford classification, and gain or loss of thigh muscle were investigated. Thigh muscle mass did not correlate with changes in apolipoproteins A1, B, fasting glucose, post-oral glucose tolerance test 2 h-blood glucose, HbA1c, and Rutherford classification. Among patients with muscle gain post-endovascular treatment, apolipoproteins A1 increased significantly, while apolipoproteins B levels were similar. Post-oral glucose tolerance test 2 h-blood glucose levels decreased. Preferable metabolic changes were observed in patients with skeletal muscle gain contrasted with muscle loss.

## Introduction

Skeletal muscles modulate lipid and glucose metabolism, while lipids and glucose regulate skeletal muscles and vice versa [1,2]. Accordingly, lipids and glucose contribute to the development and progression of atherosclerosis; thus, the gain or loss of skeletal muscle is associated with the patients’ vulnerability to cardiovascular events and all-cause mortality [3].

Using skeletal muscles to maintain physical activity [4] influences lipid metabolism [1,2]. Moreover, when receiving statins, the blood levels of apolipoprotein A1 (Apo A1) and B (Apo B) are reportedly significant predictors of myocardial infarction and all-cause mortality among patients with known coronary artery disease. Therefore, these markers may be more suitable for cardiovascular risk assessment [5]. Among atherosclerotic lipid and glucose parameters, glycated hemoglobin A1c (HbA1c) / A1 [6] and Apo A1 / B [7] have emerged as markers used to evaluate the atherosclerotic status of patients.

Among the atherosclerotic factors, glucose tolerance impairment carries a high risk of all-cause mortality and cardiovascular events [8]. Glucose tolerance is usually evaluated by measuring blood glucose levels 2 h after loading 75 g of glucose, an examination otherwise known as the oral glucose tolerance test (OGTT). The association between impaired OGTT and atherosclerosis is reportedly stronger than that between fasting blood glucose (FBG) or HbA1c level and atherosclerosis [9]. Since skeletal muscles regulate the storage and delivery of glucose [10], gain or loss of skeletal muscle may be linked to glucose metabolism.

Additionally, Apo A1 reportedly increases glucose consumption in skeletal muscles and improves glucose uptake, leading to amelioration of glucose tolerance [11,12]. In contrast, high blood glucose down-regulates apolipoprotein M expression, influencing Apo A1 levels [13,14]. Thus, Apo A1 and diabetes correlate bidirectionally, leading to a vicious cycle [15].

A previous report demonstrated that the volume of ischemic skeletal thigh muscle among patients with above-the-knee peripheral artery disease (PAD) changes before and after revascularization of occlusive iliofemoral arteries supplying thigh muscle after endovascular treatment (EVT) [16]. PAD limits blood flow during ambulation, manifesting as intermittent claudication (IC). In addition, PAD impairs the patients’ physical activity, thereby predisposing them to a sedentary lifestyle. A sedentary lifestyle is linked to further blood sugar elevation and skeletal muscle loss through the inflammatory cascade [17,18]. PAD can be treated by ameliorating IC and promoting physical activity after EVT [19]. IC is generally classified according to the Rutherford classification [20]. However, the association between metabolic parameters and skeletal muscle mass in PAD remains unclear.

This study aimed to investigate the relationships between Apo A1 and B levels, glucose metabolism, changes in symptoms according to Rutherford classification, and gain or loss of skeletal muscle mass among patients with above-the-knee PAD following EVT. The findings in this pilot study would pave the way for an enhanced understanding of the relationship between lipid and glucose metabolism in relation to skeletal muscle mass after EVT in a clinical setting.

## Materials and Methods

### Study design

This was a single-center, prospective, observational pilot study. Patients (14 with ipsilateral and eight with bilateral lesions) with consecutive lower extremity above-the-knee iliofemoral PAD with symptomatic IC classified according to the Rutherford criteria were prospectively registered between 2016 and 2020. All patients were scheduled to undergo elective EVT for iliofemoral lesions. To exclude and minimize the drug effects, prescribed drug treatment was started at least 1 month before their enrollment in the study and maintained throughout the remaining duration of the study.

Patients undergoing maintenance hemodialysis and critical ischemia were excluded from this study because these factors are related to excessive skeletal muscle loss [21,22]. The institutional review board of the Nippon Medical School Chiba Hokusoh Hospital approved this study (IRB number: 504) on April 25, 2016. The study was conducted in accordance with the tenets of the Declaration of Helsinki. Signed informed consent was obtained from all the patients. Fig 1 shows a flowchart of the study design.

**Fig 1.**
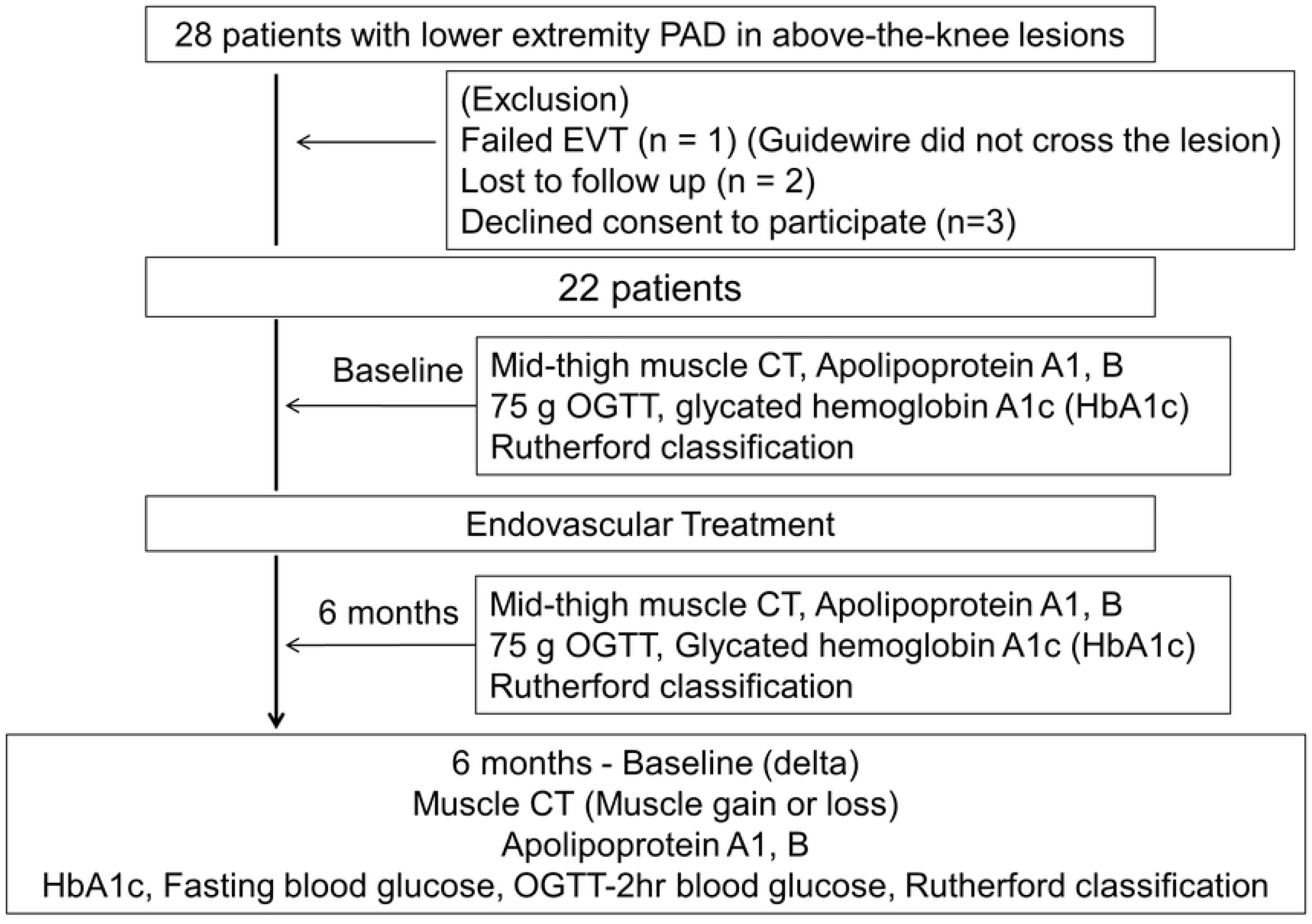
Study design flowchart.

A total of 22 patients with lower extremity PAD with iliofemoral lesions were enrolled. Blood tests, 75-g OGTTs, and computed tomography (CT) scans of the mid-thigh level of the legs were performed upon admission and at 6 months after EVT.

### Endovascular treatment

Patients intolerant to medications or contrast agents did not participate in the study. All patients were administered dual antiplatelet therapy for at least 7 days before EVT. At the start of the procedure, weight-adjusted intravenous heparin was administered with a target activated clotting time of more than 300 s. After crossing the guidewire through the target lesions, lesions were dilated using a suitable-sized balloon. The operator decided the size of the balloon. The stent diameter would exceed the reference diameter by approximately 1 mm. Post-dilatation was performed using a balloon shorter than the stent to avoid causing stent edge dissection. Stent placement was performed if the flow limitation or residual stenosis was more than 30% of the elastic recoil remaining in the target vessels. EPIC stents (Boston Scientific, Massachusetts) and SMART stents (Cordis Co, Florida) were used during the above procedure. Although efforts were made to complete the procedure with single stent placement, up to multiple stents were placed as governed by the necessity to achieve at least 30% of residual stenosis in the target lesions. Vascular patency after endovascular treatment was evaluated by the ankle-brachial index (ABI) value, which would remain throughout the study.

### Measurement of the thigh muscle area

The skeletal muscles were assessed using the cross-sectional single-slice CT, which measured the muscle volume [23]. A 64-slice CT scan, using the Toshiba Aquilion 64 system (Toshiba Medical Systems, Otawara, Japan), of the legs was performed before EVT and six months following the procedure. The cross-sectional area of the mid-thigh of both legs, between the pubic symphysis and the inferior condyle of the femur, was measured [24]. The thigh muscle area was measured using a commercial workstation (SYNAPSE; Fujifilm, Tokyo, Japan) (Fig 2). Patients were further divided into skeletal muscle gain or loss groups according to the change in the skeletal muscle area before and six months after EVT to explore the impact of muscle on metabolisms.

**Fig 2.**
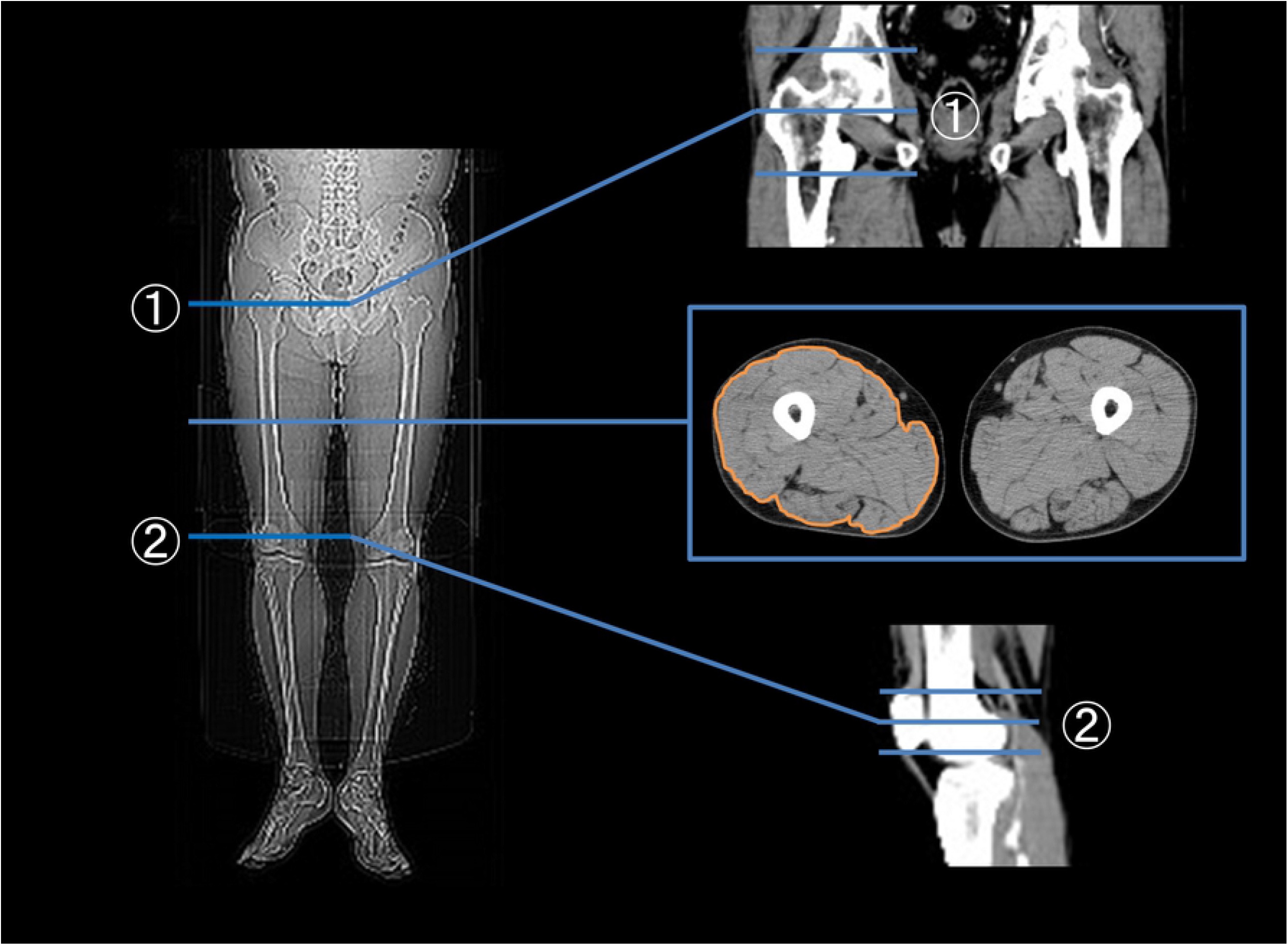
Measurement of the mid-thigh muscle area. The thigh muscle area was measured at the mid-point cross-sections of 1 and 2.① Upper edge: mid-point of the upper margin of the greater trochanter and the lower margin of the femoral condyles.② Lower edge: mid-point of the upper margin of the patella and the lower margin of the patella

### Measurement of Apo A1 and B

Serum concentrations of Apo A1 and Apo B were analyzed using immune nephelometry (Sekisui Medical Ltd., Tokyo, Japan) and measured on admission and at 6 months after EVT. The numeric values of HbA1c/Apo A1 and Apo B/Apo A1 were calculated according to the measured values.

### OGTT and glycated HbA1c

Glycated HbA1c levels (normal limit <6.5%, 47 mmol/mol) were measured to evaluate glucose control. OGTT was performed to assess glucose tolerance. OGTT was performed in the morning upon breaking an overnight fast of 8–14 h. Blood samples were taken before consuming 75 g of glucose dissolved in 250–300 mL of water and at 30 min, 60 min, and 120 min after. OGTT was performed at the time of admission and 6 months after EVT.

### ABI measurement

The ABI was measured using a Colin Wave-form analyzer (BP-203RPE III; Omron Colin, Tokyo, Japan), according to the recommendations of the American Heart Association [25]. The ABI was measured on admission and at 6 months after discharge.

### Statistical analyses

Binary or categorical data are presented as frequencies or percentages. Continuous variables are expressed as means ± standard deviations or medians with interquartile ranges (IQRs). The Shapiro–Wilk test was used to assess whether variables were normally distributed, while the chi-square test was used to assess dichotomous variables. The possible correlation analyses were performed with Spearman correlation analysis among the changes in the thigh muscle area after EVT, Apo A1 and B, fasting glucose (FBG), post 2 h-blood glucose (OGTT-2hrBG), HbA1c, Rutherford classification, and the muscle areas in the mid-thigh before and 6 months after EVT. The graphical correlation matrix displaying the positive and negative correlations between the changes in variables was obtained using the R package “Corrplot” (https://github.com/taiyun/corrplot). The concentrations of apolipoproteins and glucose, along with the metrics of atherosclerotic parameters, were compared using a single-tailed paired t-test. The PWR (Basic Functions for Power Analysis) package was used to analyze statistical power [26]. The statistical analyses were performed using the R software (ver 3.6.2; The R Foundation for Statistical Computing, Vienna, Austria). P-values less than 0.05 were considered statistically significant.

## Results

The clinical characteristics of the patients are summarized in Table 1. The mean age of the patients was 72.4 ± 7.4 years. Most patients were male (91%), and characteristics such as hypertension (82%) and dyslipidemia (86%) were prevalent. Baseline clinical symptoms were classified according to the Rutherford classification of PAD, which included the following categories: category 1 (n = 11, 50.00%), category 2 (n = 10, 45.45%), and category 3 (n = 1, 4.55%). All patients received dual antiplatelet therapy before EVT. The location and characteristics of the lesions are summarized in Table 2. There was no significant difference between gain and loss of skeletal muscle. Clinical symptoms improved after EVT, which was sustained during the study period. Six months after EVT, 82% (n = 18) of the patients were classified as category 0 and 18% (n = 4) as category 1. All patients were successfully discharged the day after EVT without complications. Vascular patency was maintained throughout the study period and verified based on the ABI value after EVT (30 ischemic lower limbs: before EVT 0.79 ± 0.14, 6 months after EVT; 1.06 ± 0.16, 14 non-ischemic lower limbs: before EVT 1.01 ± 0.10, 6 months after EVT; 1.05 ± 0.14).

**Table 1.** Baseline characteristics of the enrolled patients.

|  | All | Gain of skeletal<br>muscle | Loss of skeletal<br>muscle | P-value |
| --- | --- | --- | --- | --- |
| Number of patients | 22 | 12 | 10 |  |
| Age (years) | $72.4 \pm 7.4$ | $69.7 \pm 7.5$ | $75.6 \pm 6.2$ | 0.06 |
| Male | 20 (91) | 12 (100) | 8 (80) | N. S |
| Hypertension | 18 (82) | 10 (83) | 8 (80) | N. S |
| Dyslipidemia | 19 (86) | 10 (83) | 8 (80) | N. S |
| Diabetes | 10 (45) | 6 (50) | 4 (40) | N. S |
| HbA1c (%) | $6.1 \pm 0.7$ | $5.8 \pm 0.4^*$ | $6.5 \pm 0.8^*$ | 0.0175* |
| BMI | $24.4 \pm 3.5$ | $25.1 \pm 2.6$ | $23.5 \pm 4.4$ | N. S |
| Past smoker | 10 (45) | 6 (50) | 4 (40) | N. S |
| Rutherford classification | $1.5 \pm 0.6$ | $1.6 \pm 0.5$ | $1.5 \pm 0.7$ | N. S |
| Medications |  |  |  |  |
| Dual antiplatelet therapy | 22 (100) | 12 (100) | 10 (100) | N. S |
| Aspirin | 19 (86) | 10 (83) | 9 (90) | N. S |
| Clopidogrel | 9 (41) | 5 (42) | 4 (40) | N. S |
| Cilostazol | 6 (27) | 2 (8) | 4 (40) | N. S |
| Prasugrel | 6 (27) | 4 (33) | 2 (20) | N. S |
| Statin | 21 (95) | 11 (92) | 10 (100) | N. S |
| ACE-I / ARB | 22 (100) | 12 (100) | 10 (100) | N. S |
| β-blocker | 15 (68) | 8 (67) | 7 (70) | N. S |

Data are presented as numbers (%). p \* < 0.05, p \*\* < 0.01, Not significant (N. S). ACE-I, angiotensin-converting enzyme inhibitor; ARB, angiotensin II receptor blocker. Statin regimen is listed in Supplementary Table 1

**Table 2.** Location and characteristics of lesions.

| Variables | All | Gain of skeletal<br>muscle | Loss of skeletal<br>muscle | p-value |
| --- | --- | --- | --- | --- |
| Location of lesions | N = 30 | N = 17 | N = 13 |  |
| Unilateral lesion | 14 (64) | 7 (58) | 7 (70) | N.S |
| Bilateral lesions | 8 (36) | 5 (42) | 3 (30) |  |
| Iliac artery | 16 (53) | 10 (59) | 6 (46) | N.S |
| Superficial femoral artery | 9 (30) | 6 (35) | 5 (38) | N.S |
| Common femoral artery | 1 (3) | 0 (0) | 1 (8) | N.S |
| Popliteal artery | 3 (13) | 1 (6) | 0 (0) | N.S |
| Superficial + popliteal artery | 1 (5) | 0 (0) | 1 (8) | N.S |
| TASC II classification |  |  |  |  |
| Type A | 17 (58) | 10 (59) | 7 (54) | N.S |
| Type B | 6 (20) | 3 (18) | 3 (23) | N.S |
| Type C | 2 (6) | 0 (0) | 2 (15) | N.S |
| Type D | 5 (16) | 4 (23) | 1 (8) | N.S |
| Stent implantation | 24 (80) | 15 (88) | 9 (70) | N.S |

Data are presented as numbers (%). N.S, Not significant; TASC, Trans-Atlantic Intersociety Consensus

### Thigh muscle area before and after EVT

The thigh muscle area of each patient changed after EVT (delta muscle area: 2.5±8.1 cm^2^, n = 22). Twelve patients gained skeletal muscle (delta thigh muscle area was positive: 8.4±5.9 cm^2^), and 10 patients lost skeletal muscle (delta thigh muscle area was negative: −4.7±2.4 cm^2^) according to the change in the skeletal muscle area before and six months after EVT (Table 3).

**Table 3.** Change in muscle area, Apolipoprotein A1, Apolipoprotein B, glucose metabolism, and Rutherford classification.

| All (n=22) | Before | After 6 months | P-value | Delta value |
| --- | --- | --- | --- | --- |
| Muscle area (cm<sup>2</sup>) | 239.8 ± 34.8 | 243.1 ± 40.8 | 0.21 | 2.5 ± 8.1 |
| Apo A1 (mg/dl) | 119.0 ± 17.4 ** | 129.6 ± 19.6 ** | 0.0027 | 10.7 ± 14.7 |
| Apo B (mg/dl) | 76.3 ± 17.3 * | 81.3 ± 17.7 * | 0.0404 | 5.0 ± 9.4 |
| HbA1c (%) | 6.1 ± 0.7 | 6.2 ± 0.8 | 0.74 | 0.073 ± 0.517 |
| Apo B/Apo A1 | 0.65± 0.16 | 0.64 ± 0.16 | 0.11 | -0.012 ± 0.081 |
| HbA1c/Apo A1 | 0.053 ± 0.011 * | 0.049 ± 0.011 * | 0.0489 | 0.073 ± 0.517 |
| Fasting Blood Glucose (mg/dl) | 110.8 ± 20.4 | 116.6 ± 24.1 | 0.24 | 5.9 ± 22.5 |
| 2 hr-OGTT Blood Glucose (mg/dl) | 193.5 ± 78.9 | 182.5 ± 79.6 | 0.29 | -11.0 ± 48.0 |
| Rutherford classification | 1.5 ± 0.6 | 0.2 ± 0.4 | p < 0.001 | -1.4 ± 0.8 |

|  | Gain of skeletal muscle n = 12 |  |  | Loss of skeletal muscle n = 10 |  |  |
| --- | --- | --- | --- | --- | --- | --- |
|  | before | 6 months | P-value | before | 6 months | P-value |
| Delta muscle area (cm<sup>2</sup>) | NA | 8.41 ± 5.93 | NA | NA | -4.67±2.41 | NA |
| Apo A1 (mg/dl) | 121.8 ± 15.1 ** | 136.5 ± 19.5 ** | p < 0.001 | 115.6 ± 20.1 | 121.4 ± 17.1 | 0.32 |
| Apo B (mg/dl) | 76.4 ± 19.2 | 80.5 ± 17.0 | 0.10 | 78.6 ± 19.8 | 82.3 ± 19.3 | 0.24 |
| HbA1c (%) | 5.8 ± 0.4 | 6.0 ± 0.4 | 0.97 | 6.5 ± 0.8 | 6.5 ± 1.1 | 0.50 |
| Apo B/Apo A1 | 0.64 ± 0.17 | 0.60 ± 0.17 | 0.069 | 0.69 ± 0.19 | 0.68 ± 0.16 | 0.66 |
| HbA1c/Apo A1 | 0.049 ± 0.007 ** | 0.045 ± 0.008 ** | 0.007 | 0.058 ± 0.004 | 0.054 ± 0.004 | 0.37 |
| Fasting Blood Glucose (mg/dl) | 110.2 ± 16.8 | 111.4 ± 22.2 | 0.83 | 111.5 ± 16.8 | 122.9 ± 25.9 | 0.094 |
| 2 hr-OGTT Blood Glucose (mg/dl) | 189.7 ± 67.5 | 170.6 ± 69.7 | 0.075 | 198.1 ± 94.4 | 196.7 ± 91.8 | 0.93 |
| Rutherford classification | 1.6 ± 0.5 *** | 0.3 ± 0.5 *** | p < 0.001 | 1.5 ± 0.7 *** | 0.1 ± 0.3 *** | p < 0.001 |

p \*<0.05, p \*\*<0.01, p \*\*\* < 0.001 Not Applicable (NA).

### Correlation analysis among the change in Apo A1, B, glucose metabolism, intermittent claudication symptom, and skeletal muscle mass

The analyses were performed to investigate possible correlations among changes in the delta values of Apo A1 and B, fasting glucose, post 2 h-blood glucose, HbA1c, Rutherford classification, and delta thigh muscle area after EVT. Delta muscle area and delta value of each parameter were not significantly correlated (Figure 3). Equivalent correlation analyses were performed in muscle gain and muscle loss patients (Supplementary Figures). Among the measured parameters, delta Apo A1 correlated with delta HbA1c/Apo A1 (R = −0.76, p < 0.01) and delta B (R = −0.58, p = 0.005); delta Apo B correlated with B/A1 (R = 0.43, p=0.005); delta HbA1c correlated with HbA1c/Apo A1 (R = 0.58, p < 0.01); delta HbA1c/Apo A1 correlated with delta Apo B/A1 (R = 0.45, p = 0.038) and delta Rutherford classification (R = 0.51, p = 0.015); delta FBG correlated with delta OGTT-2hr BG (R = 0.49, p = 0.019) and Rutherford classification (R = 0.45, p = 0.034) (Fig 3).

**Fig 3.**
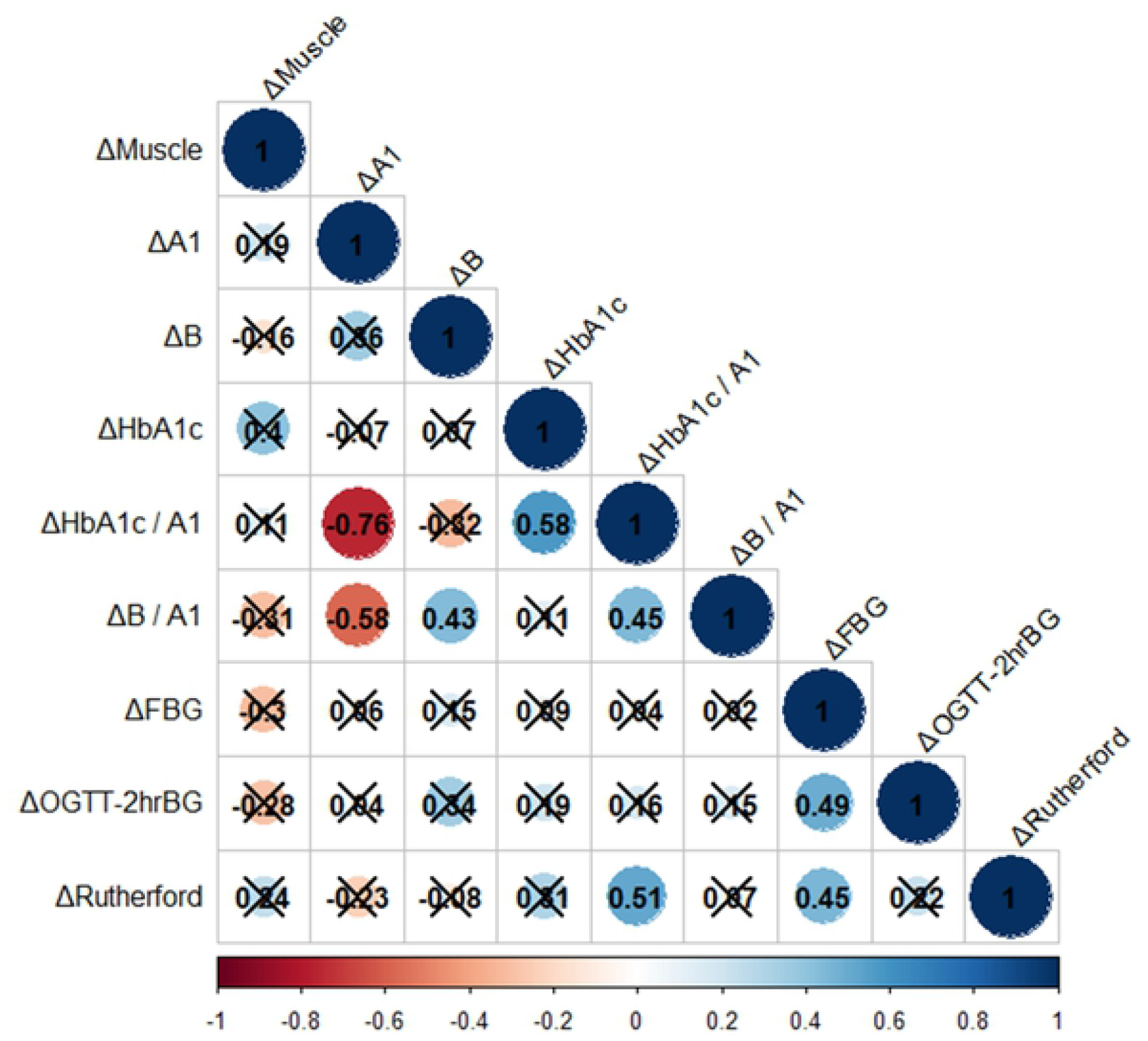
Correlation matrix between delta values of Apo A1, B, fasting glucose, post 2 h-blood glucose, HbA1c, Rutherford classification, and delta thigh muscle area after EVT.

A correlation matrix between delta values is drawn based on Spearman correlation analysis. Correlation coefficients are colored according to the value combined with significant values, and p-values ≥ 0.05 were considered insignificant and have been crossed out.

### Glucose control and glucose tolerance before and after EVT

Overall, blood glucose levels did not change while fasting (pre: 110.8 ± 20.4 mg/dL, 6 months: 116.6 ± 24.1 mg/dL) and 2 h after OGTT (pre: 193.5 ± 78.9 mg/dL; 6 months: 182.5 ± 79.6 mg/dL) (Figure 4). HbA1c did not change either (pre: 6.1 ± 0.4%, 6 months: 6.2 ± 0.4%) and was similar among patients with skeletal muscle gain (pre: 5.8 ± 0.4%, 6 months: 6.0 ± 0.4%) and loss (pre: 6.5 ± 0.8%, 6 months: 6.5 ± 1.1%) (Table 2).

**Fig 4.**
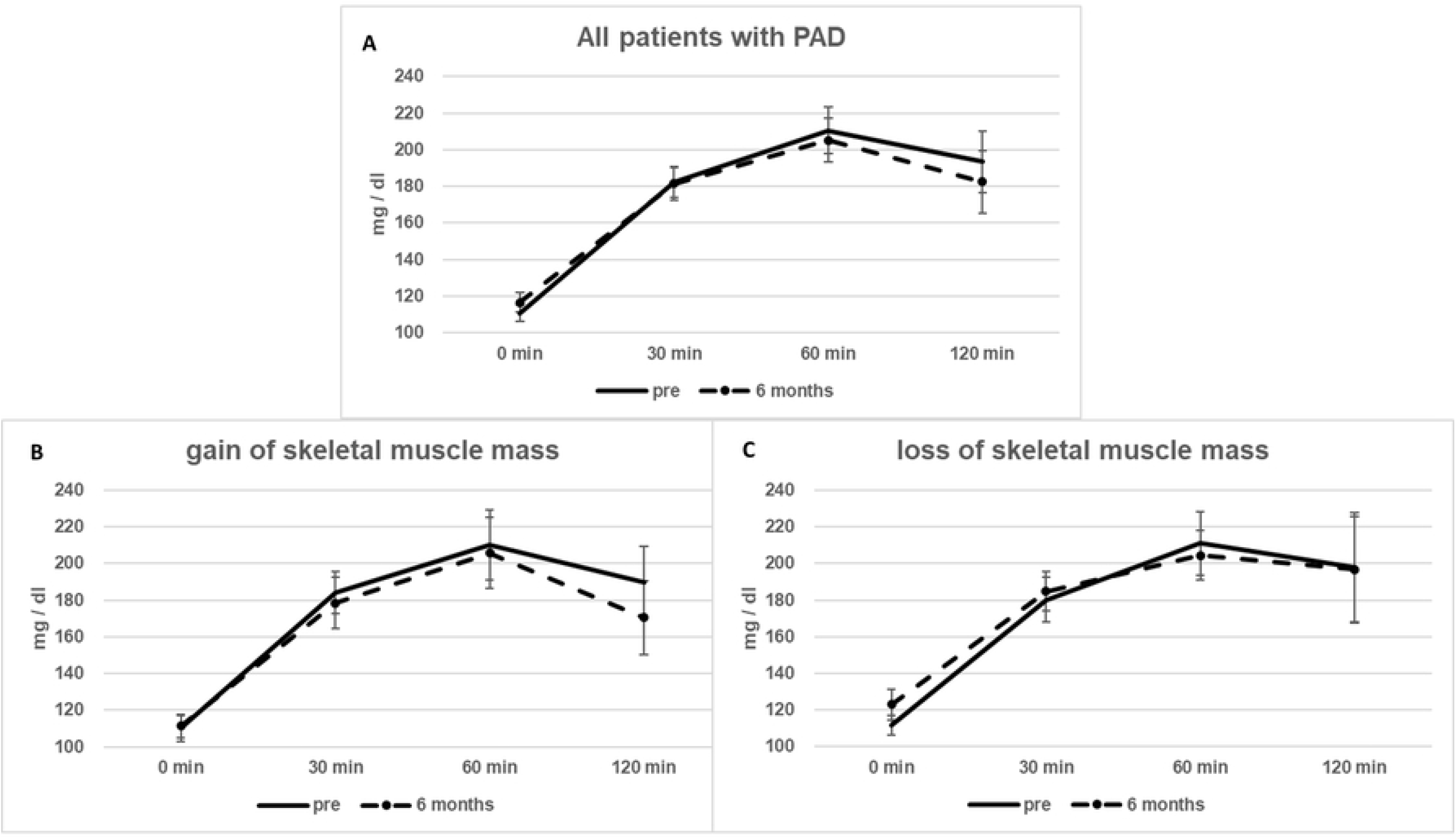
Results of the 75-g OGTT. 4A: In all the patients, the blood glucose level did not change during fasting (pre: 110.8 ± 20.4 mg/dL, 6 months: 116.6 ± 24.1 mg/dL), 30 min after OGTT (pre: 182.3 ± 38.4 mg/dL, 6 months: 181.3 ± 41.2 mg/dL), 60 min after OGTT (pre: 210.5 ± 59.5 mg/dL, 6 months: 205.2 ± 56.6 mg/dL), and 2 h after OGTT (pre: 193.5 ± 78.9 mg/dL, 6 months: 182.5 ± 79.6 mg/dL). 4B: In patients with gain of skeletal muscle, the blood glucose level did not change during fasting (pre: 110.2 ± 16.8 mg/dL, 6 months: 111.4 ± 22.2 mg/dL), 30 min after OGTT (pre: 184.2 ± 40.0 mg/dL, 6 months: 178.5 ± 47.7 mg/dL), and 60 min after OGTT (pre: 210.2 ± 65.7 mg/dL, 6 months: 205.8 ± 67.7 mg/dL) but decreased 2 h after OGTT (pre: 189.7 ± 67.5 mg/dL, 6 months: 170.6 ± 69.7 mg/dL, p = 0.0754). 4C: In patients with loss of skeletal muscle, the fasting glucose level was increased (pre: 111.5 ± 16.8 mg/dL, 6 months: 122.9 ± 25.9 mg/dL, p = 0.0939), but there was no alteration in 30 min after OGTT (pre: 180.1 ± 38.5 mg/dL, 6 months: 184.7 ± 34.1 mg/dL), 60 min after OGTT (pre: 211.0 ± 54.7 mg/dL, 6 months: 204.5 ± 43.3 mg/dL), and 2 h after OGTT (pre: 198.1 ± 94.4 mg/dl, 6 months: 196.7 ± 91.8 mg/dl).

In patients with skeletal muscle gain, the glucose level did not change during fasting (pre: 110.2 ± 25.0 mg/dL, 6 months: 111.4 ± 22.2 mg/dL) but tended to decrease 2 h after OGTT (pre: 189.7 ± 67.5 mg/dL, 6 months: 170.6 ± 69.7 mg/dL, *p* = 0.0754) (Figure 4B). In patients with loss of skeletal muscle, their fasting glucose level tended to increase (pre: 111.5 ± 16.8 mg/dl, 6 months: 122.9 ± 25.9 mg/dl, p = 0.0939), and their blood glucose levels 2 h after OGTT remained unaltered (pre: 198.1 ± 94.4 mg/dL, 6 months: 196.7 ± 91.8 mg/dL) (Figure 4C).

### Change of Apo A1 and B before and after EVT

Overall, Apo A1 levels increased after EVT (pre: 119.0 ± 17.4 mg/dL, 6 months: 129.6 ± 19.6 mg/dL, *p* = 0.0027). Apo A1 levels significantly increased in patients with skeletal muscle gain (pre: 121.8 ± 15.1 mg/dL, 6 months: 136.5 ± 19.5 mg/dL, *p* < 0.001) (Table 2). Apo B levels did not change in patients with either skeletal muscle gain (pre:76.4 ± 19.2 mg/dL, 6 months: 80.5 ± 4.9 mg/dL) or skeletal muscle loss (pre: 78.6 ± 19.8 mg/dL, 6 months: 82.3 ± 19.3 mg/dL) (Table 2).

### Atherosclerotic metrics of Apo B/Apo A1 and HbA1c/Apo A1

Overall, Apo B/Apo A1 levels (pre: 0.66 ± 0.18, 6 months: 0.64 ± 0.16) decreased in patients with skeletal muscle gain (pre: 0.64 ± 0.18, 6 months: 0.60 ± 0.17, *p* = 0.0693) but did not change significantly in those with skeletal muscle loss (pre: 0.69 ± 0.19, 6 months: 0.68 ± 0.16). In addition, HbA1c/Apo A1 significantly decreased (pre: 0.053 ± 0.011, 6 months: 0.049 ± 0.011, *p* = 0.0489) (Table 2). However, the decrease was only significant among patients with skeletal muscle gain (pre: 0.049 ± 0.007, 6 months: 0.045 ± 0.008, *p* = 0.0070) but not in those with skeletal muscle loss (pre: 0.058 ± 0.013, 6 months: 0.054 ± 0.012) (Table 2).

## Discussion

The major findings in this study are as follows: first, there was no significant correlation between change in muscle area and Apo A1 and Apo B levels, glucose metabolism, and Rutherford classification. The change in Apo A1 levels negatively correlated with HbA1c/A1 and B/A1 changes. Second, when the patients were categorized as having muscle gain or loss based on the thigh muscle CT after EVT, Apo A1 levels were significantly increased in patients with skeletal muscle gain after EVT. Additionally, Apo B levels did not change in patients with skeletal muscle gain or loss. Third, glucose tolerance improved muscle gain, while fasting glucose levels decreased in patients with muscle loss.

### Changes in Apo A1 and B, glucose metabolism, intermittent claudication symptom, and skeletal muscle mass

While there was no significant correlation between delta muscle area and delta values of Apo A1 and B, glucose metabolisms, and Rutherford classification, Apo A1 levels were significantly increased in patients with skeletal muscle gain. Since exercise would increase Apo A1 levels [27], successful EVT can promote physical activity in patients with PAD, which is associated with the increase in Apo A1 levels in the present study. While no specific exercise was implemented in this study, only a modest change in physical activity of walking after EVT can modify the metabolism to anti-atherosclerotic [28,29]. Apo B levels did not change significantly in patients with skeletal muscle gain or loss since all the enrolled patients with PAD underwent statin therapy, which might explain why there was no change.

According to the current clinical practice guidelines, initiation of statin therapy is recommended to prevent subsequent cardiovascular events in established cardiovascular disorders [30]. The efficacy of statin therapy has been well established in numerous studies. The statin on-treatment blood level of Apo A1 and Apo B were significant predictors of myocardial infarction and all-cause mortality in patients with known coronary artery disease and might be more suitable for assessing cardiovascular risk [5]. Apo A1 functions as a major component of the high-density lipoprotein complex to clear cholesterol from white blood cells inside the artery walls [31]. WBCs are less likely to become lipid overloaded, transform into foam cells, and die, contributing to the progression of atherosclerosis [31]. Additionally, among patients treated with statins, an increase in Apo A1 levels was associated with a reduced risk of major cardiovascular events, while high-density lipoprotein (HDL) cholesterol was not [32]. Conversely, Apo B particles are atherogenic and entrapped within the arterial walls. They initiate and cause persistent inflammation, thus resulting in advanced atherosclerosis [33].

We previously demonstrated that skeletal muscle mass increases after EVT, specifically in patients with normoglycemic control (HbA1c < 6.5) before EVT [34]. Thus, if the PAD patients were grouped based on skeletal muscle volume gain or loss, patients with skeletal muscle gain would be considered closer to normoglycemia than those with loss of skeletal muscle. HbA1c is clinically used to monitor glycemic control among diabetic patients [35]. None of the patients in the present study were obliged to cut caloric intake or seek dietary guidance, as apparent from the unchanged HbA1c values [36]. Glucose tolerance was measured among enrolled patients with 75-g OGTT. Blood glucose levels 2 h after 75-g OGTT have been associated with atherosclerosis more than fasting glucose or HbA1c levels [9]. Blood glucose is regulated by glucose transporters (GLUTs). Among glucose transporters, GLUT4 plays a central role in glucose homeostasis [37]. In rodent models, target disruption of glucose transporters in muscles causes insulin resistance and glucose intolerance [38]. Additionally, glucose transporters in the thigh muscle are upregulated by exercise, which improves glucose tolerance [39]. Therefore, muscles function as organs that control serum glucose levels. In the present study, there was no correlation between the change in skeletal muscle volume and glucose metabolism; however, PAD patients with skeletal muscle gain tended to show improved glucose tolerance, while those with skeletal muscle loss had a deteriorated fasting glucose level. Additionally, while Apo A1 improves glucose tolerance [11,12], high blood glucose levels downregulate Apo A1 expression. In patients with skeletal muscle gain, Apo A1 levels increased significantly, and glucose tolerance tended to improve, while blood glucose levels were high in patients with muscle loss. The results of this study are consistent with those of previous reports [11,12].

### Metrics of atherosclerosis: HbA1c/Apo A1 and Apo B/Apo A1

Apolipoproteins may be atherogenic or anti-atherogenic and serve as biological markers. A recent study has shown that the ratio of HbA1c/A1 after myocardial infarction helps predict all-cause mortality and major adverse cardiac events, including nonfatal myocardial infarction, cardiac death, and revascularization [6]. In addition, numerous studies have demonstrated that the Apo B/Apo A1 ratio can be used as a parameter of atherosclerotic conditions [5]. Recent studies have shown that Apo B/Apo A1 are inadequate to assess the recurrent risk of cardiovascular events among on-statin-treated patients [40,41]. In the present study, although there was no significant correlation between changes in skeletal muscle and delta HbA1c/Apo A1 and Apo B/Apo A1, delta Apo A1 significantly correlated with delta HbA1c/Apo A1 and Apo B/Apo A1. Accordingly, Apo A1 modified the metrics of atherosclerosis (HbA1c/Apo A1 and Apo B/Apo A1). Thus, the change in Apo A1 after EVT would impact atherosclerotic conditions. Additionally, HbA1c/A1 was significantly decreased among patients with muscle gain after EVT. However, the previous studies lacked a consecutive long-term follow-up [5,6], and long-term changes should be further studied.

### Study limitations

This study has several limitations. First, even though the calculated statistical power was adequate in this preliminary study, recruitment of a larger number of participants estimated based on the data from this pilot study would strengthen the results further. Second, the observational period was six months, which was relatively short. Observation for a more extended period is needed to assess whether the multifaceted favorable effect of EVT continues. Third, we recruited patients with PAD with mild claudication and excluded patients with below-the-knee lesions, hemodialysis treatment, and critical limb ischemia. Recent research has revealed that patients with PAD are more likely diabetic, frequently undergoing hemodialysis treatment, and mostly older than patients with coronary artery disease, suggesting that patients with PAD visit clinicians only when cardiovascular pathology has already advanced [42]. The early detection of PAD, as in this study, can confer significant benefits and delay atherosclerotic progression [43]. Fourth, we did not recruit patients with neuropathic muscle atrophy, which was common in patients with diabetic neuropathy [44] or lumbar disc herniation [45], as their presentations could partially overlap with PAD. Including patients with typical IC would exclude such subsets of patients. Fifth, we did not implement duplex ultrasound assessment on vascular patency, which represents a more accurate diagnostic test than resting ABI assessment. Sixth, we did not implement a specific exercise regimen among the PAD cohort due to institutional limitations. The effect of supervised exercise after EVT will be explored in a future study. Finally, we did not objectively evaluate the patient’s normal and subsequent physical activity levels. Levels of physical activity are associated with changes in metabolism after EVT, which were only based on the Rutherford criteria in this study and were derived from subjective symptoms. To date, physical activity assessment is largely derived from questionnaires and interviews [46, 47]. Capturing and incorporating the patients’ objective baseline physical activity and changes during the study with new technology such as wearable monitoring devices [48] would reveal the confounding factor influencing the impact of EVT on the metabolism of patients with PAD in the upcoming clinical trials [49].

## Conclusions

This study revealed the unidentified relationships between metabolic parameters and skeletal muscle mass in clinical settings. EVT had a positive impact on apolipoproteins and glucose metabolisms in patients with PAD. Notably, in patients with skeletal muscle gain after EVT, an increase in anti-atherosclerotic apolipoprotein Apo A1 levels and possible improvement in glucose tolerance are expected. Extended studies are needed to fill the knowledge gap regarding the regulation of metabolism and skeletal muscle mass.

## Data Availability

Data used in this paper cannot be shared publicly because of its confidential nature and to prevent deductive disclosure of the identity of the study participants.

## Acknowledgments

We would like to express our overflowing gratitude to all the staff of Nippon Medical School, Chiba Hokusoh Hospital Cardiovascular Centre, for their valuable therapeutic and technical support.

## Author Contributions

TI: HK: conception and design, analysis, interpretation of data, and drafting of manuscript. TM, MT, MM, NK, KA, MM, HT, WS, and YM: conception and design, critical revision of manuscript. All authors have read and approved the final manuscript.

## Supporting Information

**S1 Fig. The correlational matrix between delta values of Apo A1, B, fasting glucose, post 2 h-blood glucose, HbA1c, Rutherford classification, and delta thigh muscle area after EVT in skeletal muscle gain patients**.

**S2 Fig. The correlational matrix between delta values of Apo A1, B, fasting glucose, post 2 h-blood glucose, HbA1c, Rutherford classification, and delta thigh muscle area after EVT in skeletal muscle loss patients**.

**S1 Table. Statin regimen of the study cohort**.

